# After the Infection: A Survey of Pathogens and Non-communicable Human Disease

**DOI:** 10.1101/2023.09.14.23295428

**Authors:** Michael Lape, Daniel Schnell, Sreeja Parameswaran, Kevin Ernst, Shannon O’Connor, Nathan Salomonis, Lisa J. Martin, Brett M. Harnett, Leah C. Kottyan, Matthew T. Weirauch

## Abstract

There are many well-established relationships between pathogens and human disease, but far fewer when focusing on non-communicable diseases (NCDs). We leverage data from The UK Biobank and TriNetX to perform a systematic survey across 20 pathogens and 426 diseases, primarily NCDs. To this end, we assess the association between disease status and infection history proxies. We identify 206 pathogen-disease pairs that replicate in both cohorts. We replicate many established relationships, including *Helicobacter pylori* with several gastroenterological diseases and connections between Epstein-Barr virus with multiple sclerosis and lupus. Overall, our approach identified evidence of association for 15 pathogens and 96 distinct diseases, including a currently controversial link between human cytomegalovirus (CMV) and ulcerative colitis (UC).

We validate this connection through two orthogonal analyses, revealing increased CMV gene expression in UC patients and enrichment for UC genetic risk signal near human genes that have altered expression upon CMV infection. Collectively, these results form a foundation for future investigations into mechanistic roles played by pathogens in NCDs. All results are easily accessible on our website, https://tf.cchmc.org/pathogen-disease.

## Introduction

Humans are exposed to infectious agents throughout life. Many of the communicable diseases associated with specific infectious agents are well-characterized^1^. In acute infection, virus-driven mechanisms are clearly and predominantly seen as the agent of disease. For example, respiratory syncytial virus (RSV) causes an upper-respiratory disease that can be life-threatening in young infants and causes cold-like symptoms in adolescents and adults^2^. Likewise, varicella zoster virus (VZV) causes varicella (chickenpox) upon primary infection. This infection typically occurs during childhood and is well tolerated. However, if primary infection occurs in infants, adults, or the immunocompromised, the viral infection is less well contained, and the virally mediated pathology can be life-threatening^3^. Even after primary infection, VZV lies dormant for decades, reactivating in a portion of adults with the lytic virus directly leading to zoster (shingles)^3^.

The role of infectious agents in non-communicable diseases (NCDs) is much less well-explored, although several associations are well-known. For example, *Helicobacter pylori* infection is the strongest risk factor for gastric cancers^4^. Likewise, multiple studies have demonstrated an important role for both hepatitis B (HBV) and C viruses (HCV) in the development of cirrhosis and other chronic liver diseases^5,6^. Further, certain strains of human papillomavirus (HPV) are known to cause a large proportion of cervical cancers and a smaller proportion of several other cancers^7^. Each of these pathogens, as well as several others, have been classified as biological carcinogens by the International Agency for Research on Cancer (IARC)^8^. Indeed, it is estimated that up to 15% of all new cancer diagnoses are attributable to these and other infectious agents^9^. More recently, strong epidemiologic and mechanistic links have been identified between Epstein-Barr virus (EBV) infection and multiple sclerosis (MS)^10–12^, following decades of suggestive epidemiological and molecular evidence^13,14^. Many unknown pathogen-disease connections likely remain yet to be discovered.

NCDs are often chronic diseases and have complex etiologies. Unlike acute viral infections, the virus itself might not contribute to the full etiology of virally associated NCDs. While many individuals might be exposed and mount an immune response to a virus, virally associated NCDs often occur only in a small subset of individuals in the context of specific genetic factors and other environmental exposures^15^.

Historically, connections between pathogens and diseases have been made one pair at a time. However, the recent establishment of large-scale national biobanks and the general shift to electronic health records (EHRs) enables the concurrent analysis of many pathogen-disease pairs. Here, we leverage biobank data from The UK Biobank (UKB) and EHR data from TriNetX, LLC (TNX), resources containing both serologic and diagnostic records, enabling the systematic detection of associations between multiple pathogens and multiple diseases simultaneously. Using a discovery-replication approach, our analysis reveals 206 replicated pathogen-disease associations, including previously established pathogen-disease links and many novel or previously suggestive relationships. In particular, we identify strong evidence for the currently controversial connection between cytomegalovirus (CMV) infection and ulcerative colitis (UC).

Orthogonal analyses of this relationship reveal that: (1) patients with UC have elevated CMV mRNA levels in intestinal tissue samples compared to healthy controls, and (2) UC genetic risk loci are enriched near human genes that change expression upon CMV infection in two independent datasets. Collectively, these results implicate multiple pathogens in dozens of non-communicable human diseases, providing a unique and powerful resource for future studies of the mechanistic roles played by these pathogens in disease development.

## Results

### Pathogen-disease data collection in two large independent cohorts

We employed a discovery-replication experimental design using two newly available resources, The UK Biobank (UKB) and TriNetX (TNX). For our discovery cohort, we used the portion of UKB participants that had 45 antibody titers systematically measured, representing immune responses to 20 unique pathogens (9,429 UKB participants). Within the set of UKB subjects with serologic data, setting aside International Classification of Diseases 10^th^ revision (ICD10) codes A00 - B99 covering “Certain infectious and parasitic diseases”, we found 399 non-communicable diseases (NCDs) and 27 communicable diseases (ICD10 codes between C00 and O99) for which we were statistically powered to test (**Supplemental Dataset 1**). The primary focus of this study is on the 399 NCDs with a secondary assessment of communicable diseases that are associated with non-causative pathogens. This discovery cohort (UKB) has the advantage of a carefully controlled experimental design with many covariates for the detection of confounding effects, but it has the disadvantage of a relatively small number of subjects.

We used data extracted from The TriNetX Research Network as our independent replication cohort. These data contain both the clinical diagnoses and the serologic test results required for our model for over 11 million individuals. A total of 209 separate binary (positive/negative) clinical laboratory tests targeting the 20 UKB pathogens were identified in the medical records provided by TNX. Utilizing the same statistical power requirements, TNX was powered to test nearly 75% of the 8,616 tested UKB pathogen-disease pairs. This replication cohort (TNX) has the disadvantage of a less well-controlled study design involving data from different clinical tests and sites, with the advantage of a very large number of subjects. The larger number of subjects enabled the use of an additional restriction requiring infection status to appear in a participant’s medical record prior to disease diagnosis (see Methods).

### Development of a statistical model with high sensitivity and specificity

We developed a workflow for systematically detecting and replicating pathogen-disease pairs within our discovery and replication cohorts (**Figure 1**). To this end, we used a logistic regression model to test for association between one of the 426 disease statuses of interest and a proxy for a history of infection by a given pathogen: either one of the 45 continuous antibody titer values (UKB) or binary clinical laboratory test results (TNX). To mitigate potential confounding, we adjusted each model for any of ten additional sociodemographic and health-related variables (**Table S1**) that were found to be significantly associated with both disease status and the pathogen proxy in the discovery cohort (**Table S2**). To prevent overfitting, a backward elimination procedure was used to prune non-significant covariates before fitting the final model (see Methods). Six infectious diseases (ICD10 codes between A00 - B99) were also included as part of a control set. Within the resulting 19,289 separate antibody-disease tests, we collapsed instances of multiple antibodies to the same pathogen by selecting the antibody with the most significant association. In total, this procedure resulted in 8,616 pathogen-disease tests.

**Figure 1.**
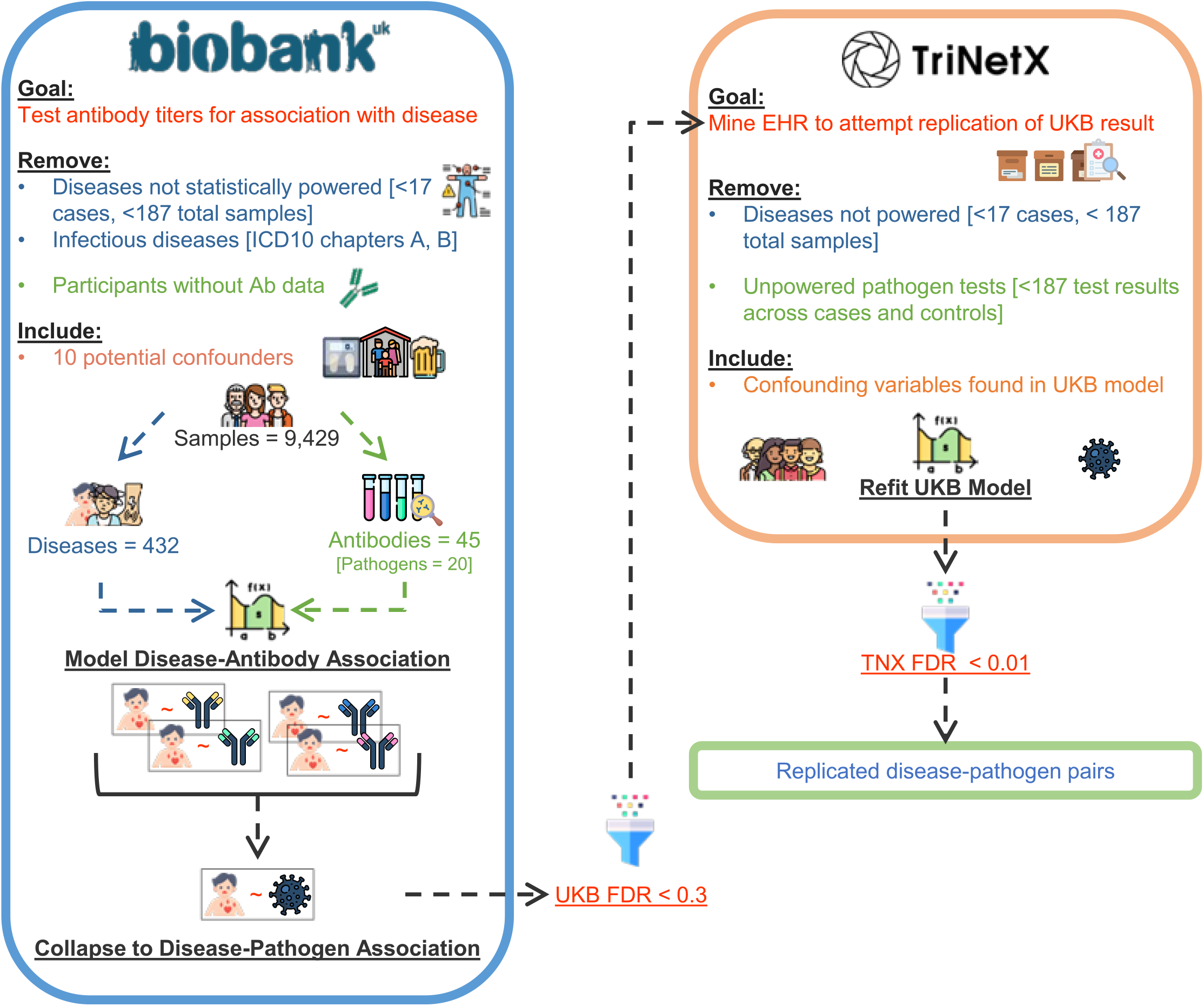
| Overview of study design. The UK Biobank (UKB) was used as the discovery cohort (left). 426 diseases with sufficient sample counts in UKB (along with positive and negative controls) were tested for association with the 45 antibody titers representing immune responses to 20 unique pathogens across 9,429 UKB participants. Models were adjusted for any of ten additional health-related and sociodemographic variables that were determined to be confounding for a particular antibody-disease pair. Significant pathogen-disease pairs identified in UKB were tested in the independent TriNetX (TNX) cohort (right). Pathogen-disease pairs that were significant in both the discovery and the replication cohort were considered replicated pairs.

The most commonly adjusted-for covariates were age, sex, and body mass index (BMI), included in 35.5%, 31.4%, and 24.4% of the UKB antibody-disease models, respectively (**Figure S1**). As expected, age was most significant in models for diseases that are more common in geriatric patients, such as “disorders of lipoprotein metabolism and other lipidaemias”, “other arthrosis”, and “other cataract”. Likewise, the strongest BMI associations were seen in models for metabolic syndrome diseases such as “obesity”, “essential primary hypertension”, and “diabetes mellitus”. Roughly a quarter of models did not require adjustment for any covariates.

We confirmed the robustness of our analytical model in our UKB discovery cohort using a permutation-based approach. To this end, we calculated an empirical p-value for each antibody-disease pair by permuting disease status across individuals (see Methods).

We observed exceptionally strong correlation (Pearson’s r > 0.99) between the nominal p-value obtained from our analytical model and the empirically derived permutation-based p-value (**Figure S2**).

To reduce the multiple testing burden, we only tested pathogen-disease pairs in the replication cohort that were statistically significant in the discovery cohort. During replication, we refit the UKB model using the replication cohort data, adjusting for the same covariates when data were available. To reduce false negatives during discovery, a lenient per-disease Benjamini-Hochberg (BH) false discovery rate (FDR) threshold of 0.3 was applied to the discovery cohort results (see Methods). Then, to reduce false positives during replication, a more stringent per-disease FDR threshold of 0.01 was used in the replication stage. Only pairs with significant associations and effects (odds ratios) in the same direction across both cohorts were considered replicated pathogen-disease relationships.

We first assessed the sensitivity of our model on a set of positive controls. “Tier 1” positive controls were identified as infectious disease diagnoses paired with their causal pathogen, e.g., hepatitis C (HCV) paired with a diagnosis of “unspecified viral hepatitis” (ICD10: B19). As expected, our model found significant associations between the disease “unspecified viral hepatitis” and both hepatitis B (HBV) and HCV pathogens. It also identified an association with the BK virus (BKV) at the more lenient discovery cohort threshold. However, upon testing for replication, only the HBV and HCV results remained significant (**Figure S3**). To assess the specificity of the model, we used a set of “Expected Negatives”, which we identified as the complement of the Tier 1 positive controls, i.e., an infectious disease diagnoses with a pathogen that does not cause the disease, such as “unspecified viral hepatitis” with Epstein-Barr virus (EBV). We excluded human immunodeficiency virus (HIV) from the Expected Negative set due to its indirect involvement in many immune-mediated diseases.

Overall, our model identified significant association for all eight (100%) of the Tier 1 positive control pathogen-disease pairs in the UKB cohort, and all eight replicated in TNX (**Table 1**). Conversely, the model identified only five (5.68%) of the Expected Negatives as significant in UKB, only one of which replicated in TNX, a diagnosis of “infectious mononucleosis” (ICD10: B27) and human herpesvirus 6 (HHV-6). Upon further investigation, this replicated Expected Negative pair represents a previously established relationship: HHV-6 infection can account for up to five percent of infectious mononucleosis (IM)-like syndrome diagnoses in adult patients^16^. Considering only the fully replicated pairs as predicted positives, and those pairs that were either identified as not significant in UKB or failed replication as predicted negatives, our model has a sensitivity of 1.0, a specificity of 0.80, and a precision of 0.89.

**Table 1.** | Results for all Tier 1 pathogen-disease pairs and other discussed pairs. The disease name with its 3-character International Classification of Diseases 10^th^ revision (ICD10) code is listed next to the paired pathogen, followed by the electronic health record statistical analysis results, including the UK Biobank (UKB) per-disease false discovery rate (FDR) and odds ratio (OR), as well as the TriNetX (TNX) per- disease FDR and OR. All eight of the Tier 1 pathogen-disease pairs and any other pairs discussed are listed. Column abbreviations: Path: Pathogen; Group: Control group (Tier 1 control, Tier 2 control, or previously Unknown association (Unk)). Disease name abbreviations: Infect: Infection; Unspec: Unspecified; Mono: Mononucleosis; Dis: Disease; GERD: Gastroesophageal reflux disease. Replication Status (Rep Status) abbreviations: REP: Replicated (UKB per-disease FDR &lt; 0.3 AND TNX per-disease FDR &lt; 0.01); DNAR: Did not attempt replication (UKB per-disease FDR > 0.3).

| Disease Name | ICD10 | Path | Group | UKB FDR | TNX FDR | UKB OR | TNX OR | Rep Status |
| --- | --- | --- | --- | --- | --- | --- | --- | --- |
| Anogenital Herpesviral Infect | A60 | HSV-2 | Tier 1 | 1.4E-03 | 2.6E-298 | 4.10 | 11.76 | REP |
| Herpesviral Infect | B00 | HSV-1 | Tier 1 | 9.5E-04 | 8.9E-266 | 2.08 | 2.93 | REP |
| Herpesviral Infect | B00 | HSV-2 | Tier 1 | 2.2E-01 | 1.6E-298 | 1.36 | 5.18 | REP |
| Herpes Zoster | B02 | VZV | Tier 1 | 6.4E-02 | 4.1E-259 | 1.43 | 14.43 | REP |
| Unspec Viral Hepatitis | B19 | HBV | Tier 1 | 2.4E-04 | 1.9E-299 | 2.28 | 7.20 | REP |
| Unspec Viral Hepatitis | B19 | HCV | Tier 1 | 6.7E-06 | 1.9E-299 | 2.96 | 11.57 | REP |
| Unspec HIV | B24 | HIV | Tier 1 | 1.3E-06 | 8.0E-300 | 38.21 | 635.37 | REP |
| Infectious Mono | B27 | EBV | Tier 1 | 5.0E-02 | 8.2E-212 | 2.14 | 8.39 | REP |
| Iron Deficiency Anemia | D50 | *H. pylori* | Tier 2 | 9.9E-03 | 1.1E-08 | 1.17 | 1.24 | REP |
| Multiple Sclerosis | G35 | EBV | Tier 2 | 2.5E-01 | 2.5E-03 | 2.55 | 4.45 | REP |
| Duodenal Ulcer | K26 | *H. pylori* | Tier 2 | 1.3E-03 | 1.7E-04 | 1.73 | 1.43 | REP |
| Peptic Ulcer Site Unspec | K27 | *H. pylori* | Tier 2 | 9.0E-02 | 5.6E-138 | 1.60 | 3.68 | REP |
| Gastritis and Duodenitis | K29 | *H. pylori* | Tier 2 | 1.5E-01 | 7.9E-134 | 1.13 | 1.97 | REP |
| Fibrosis and Cirrhosis of Liver | K74 | HCV | Tier 2 | 7.3E-02 | 6.7E-299 | 2.69 | 4.31 | REP |
| Other Dis of Liver | K76 | HBV | Tier 2 | 1.6E-02 | 7.8E-299 | 1.44 | 2.97 | REP |
| Other Dis of Liver | K76 | HCV | Tier 2 | 1.6E-02 | 7.8E-299 | 1.47 | 2.03 | REP |
| Asthma | J45 | *H. pylori* | Unk | 1.8E-02 | 2.8E-04 | 0.89 | 0.80 | REP |
| GERD | K21 | *H. pylori* | Unk | 2.1E-02 | 2.9E-07 | 0.88 | 0.66 | REP |
| Other Dis of Esophagus | K22 | *H. pylori* | Unk | 5.1E-03 | 1.4E-09 | 0.68 | 0.72 | REP |
| Diaphragmatic Hernia | K44 | *H. pylori* | Unk | 2.7E-01 | 1.4E-06 | 0.90 | 0.76 | REP |
| Inflammatory Bowel Dis | K58 | *H. pylori* | Unk | 1.9E-01 | 6.10E-29 | 0.91 | 0.48 | REP |
| Crohn's Dis | K50 | CMV | Unk | 8.7E-01 | - | 0.86 | - | DNAR |
| Crohn's Dis | K50 | EBV | Unk | 8.7E-01 | - | 1.65 | - | DNAR |
| Ulcerative Colitis | K51 | CMV | Unk | 2.0E-01 | 8.6E-06 | 1.36 | 2.78 | REP |
| Ulcerative Colitis | K51 | EBV | Unk | 3.7E-01 | - | 0.80 | - | DNAR |
| Unspec Noninfective Gastroenteritis/Colitis | K52 | CMV | Unk | 8.4E-01 | - | 0.97 | - | DNAR |
| Unspec Noninfective Gastroenteritis/Colitis | K52 | EBV | Unk | 8.4E-01 | - | 1.04 | - | DNAR |
| Systemic Lupus Erythematosus | M32 | EBV | Tier 2 | 1.8E-01 | 1.4E-21 | 3.98 | 4.96 | REP |
| Systemic Lupus Erythematosus | M32 | CMV | Unk | 4.0E-01 | - | 1.89 | - | DNAR |

We next assessed a set of 83 pathogen-NCD pairs with suggestive literature evidence, the “Tier 2” positive controls, collected via a semi-automated literature search approach (see Methods). Sixteen (19.28%) of these Tier 2 pairs were significantly associated in UKB, and 11 of the 15 with available data (68.75%) replicated in TNX. Included in these are well-known associations such as *H. pylori* with several gastroenterological diseases such as “duodenal ulcer”, “peptic ulcer, site unspecified”, and “gastritis and duodenitis”^17–19^. We also replicated connections between particular pathogens and hepatic diseases, such as “fibrosis and cirrhosis of liver” with HCV and “other diseases of liver” with both HBV and HCV^20^. Finally, our model validated the now well-established associations of EBV with multiple sclerosis (MS)^10,12,21^ and with systemic lupus erythematosus (SLE)^22,23^. Taken together, these results establish that our approach can capture both well-established and suggestive pathogen-disease relationships while maintaining a substantial degree of specificity.

### Identification of 206 replicated pathogen-disease relationships

Encouraged by the performance of our model on our positive and negative control sets, we next sought to identify novel pathogen-disease relationships. In total, of the 8,437 “unknown” (non-Tier 1 or Tier 2) pathogen-disease tests that met our requirements in UKB, 569 were significant. 462 of these pairs had sufficient data in the TNX cohort to test for replication, and 195 of these were replicated (2.3% of the total 8,437 “unknown” pairs that were initially tested) (**Figure 2a**). The 195 replicated pairs represent a diverse collection of diseases and pathogens, with 15 of the 20 tested pathogens connected to at least one of 96 distinct diseases, 89 of which are NCDs (**Figure 3** and **Supplemental Dataset 2**). Altogether, the 11 Tier 2 and the 195 “unknown” pathogen-disease relationships equate to 206 replicated associations. While these relationships are correlations, due to the size of the TNX cohort, we could restrict our study population to only those with a pathogen test result (positive or negative) before disease development while remaining statistically powered. Thus, these “temporal correlations” provide stronger evidence that the pathogen plays a role in disease development than a simple correlation. In contrast, the antibody titer data present in the UKB were obtained from the near-simultaneous measurement of all 45 antibody titers across all 9,429 pilot project participants. Thus, they cannot provide the same temporal correlations.

**Figure 2.**
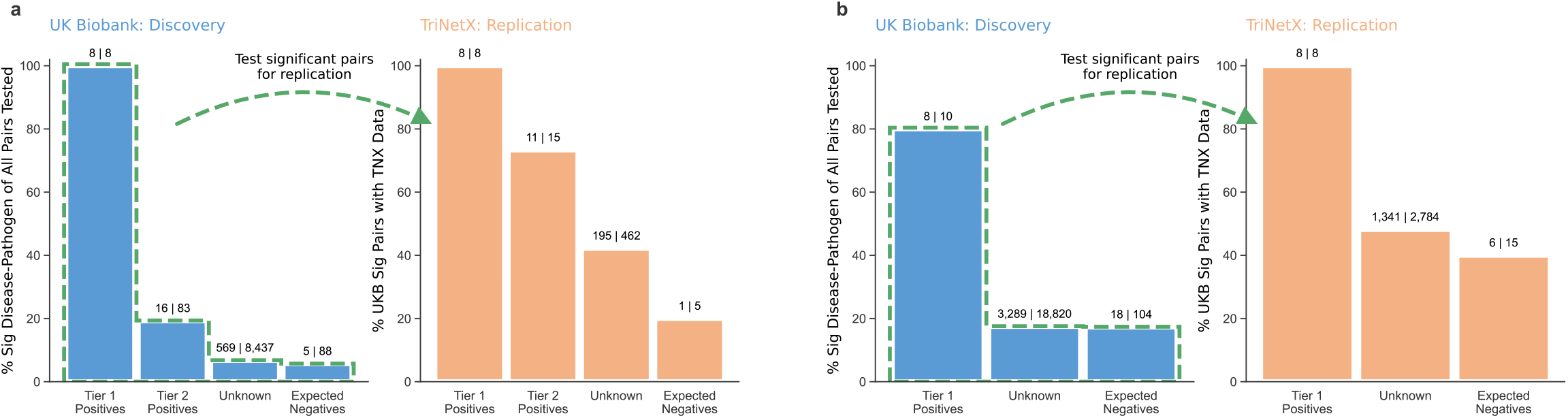
| Summary of pathogen-disease pairs identified across both cohorts. Bar charts show the percent of pathogen-disease pairs in each subgroup (“Tier 1 Positives”, “Tier 2 Positives” (ICD10 analysis only), “Expected Negatives”, and “Unknown”) that were significant in the discovery cohort (blue bars) and replication cohort (orange bars). The numbers used to calculate the percentages are indicated above each bar. All discovery cohort (blue bars) significant pairs were assessed for replication in the independent replication cohort (orange bars). Note that the total number of pairs tested for replication may not match the number of significant UK Biobank pairs due to insufficient data available in the replication cohort for some pairs. Such cases are not considered replication successes or failures. **a.** Results using International Classification of Diseases 10^th^ revision (ICD10) codes. **b.** Results using Phecodes.

**Figure 3.**
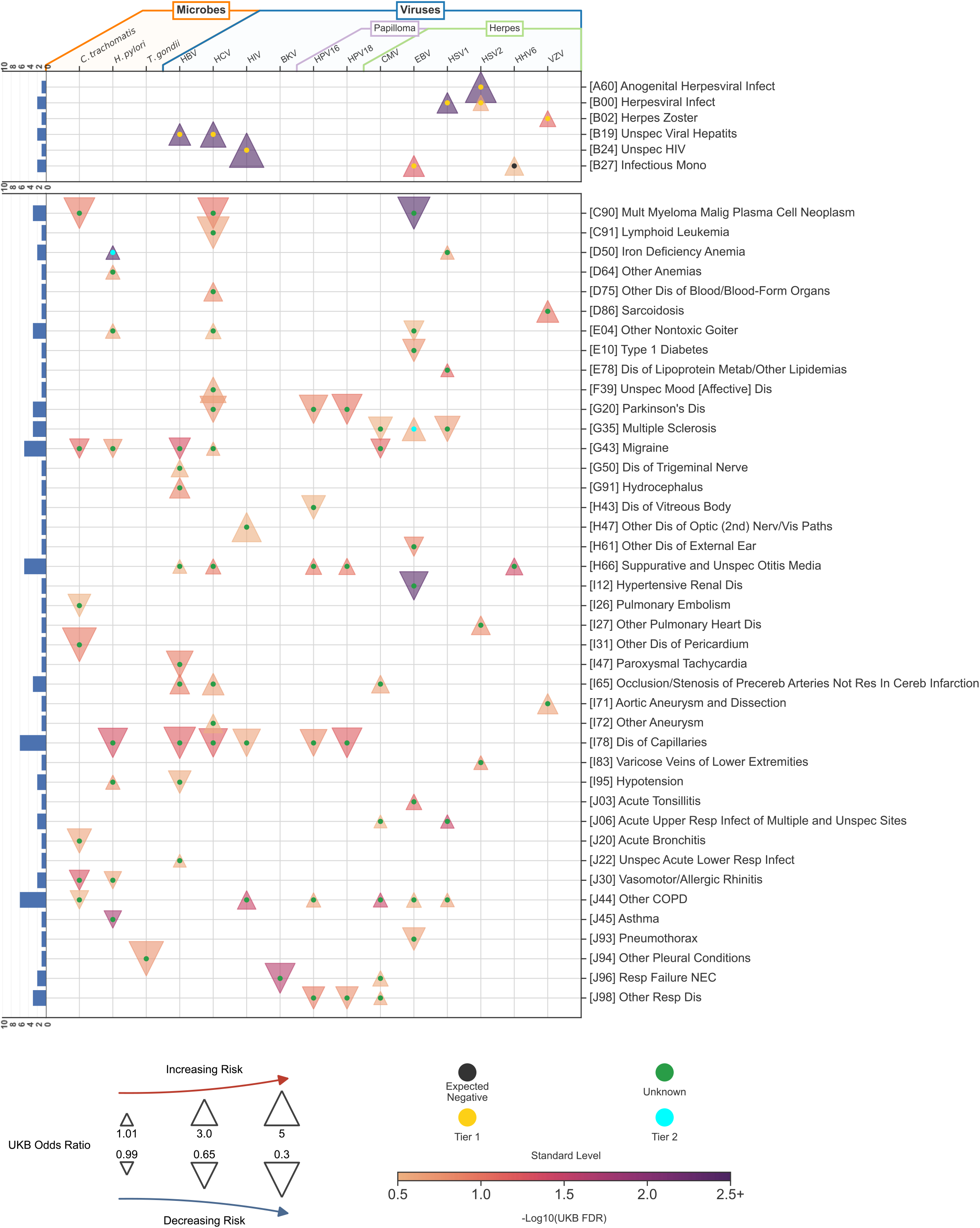
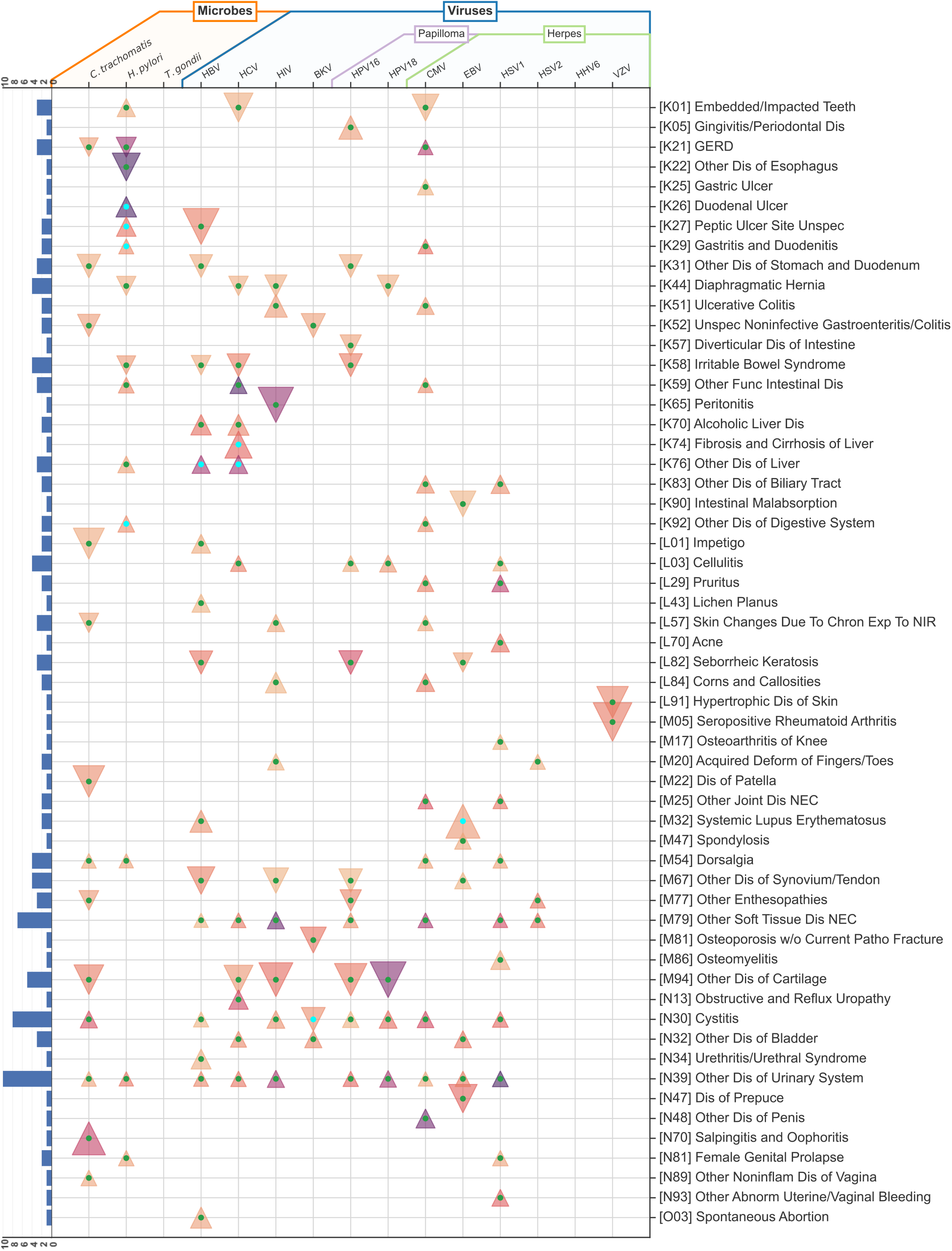
| Overview of all replicated pathogen-disease pairs. Heatmap containing all pathogens and diseases with at least one replicated result. Pathogens are first grouped by type (microbe or virus) and then sub-grouped by family if more than one member is present. Diseases are ordered by International Classification of Diseases 10^th^ revision (ICD10) code, with the Tier 1 positive controls at the top. A histogram opposite to each disease indicates the total number of replicated associations between that disease and all pathogens investigated. Each replicated result for a pathogen-disease pair is represented by a triangle either pointing up (indicating an odds ratio greater than one) or down (indicating an odds ratio less than one). The size of each triangle represents the discovery cohort (UK Biobank, UKB) odds ratio (OR) and is capped at a maximum of 5 to account for the very large effect size between “unspecified hiv disease” and human immunodeficiency virus (HIV) (OR: 38.2). The color of each triangle represents the UKB negative log base-10 corrected p-value, capped at a maximum of 2.5 to enable better visual distinction in the region covering all but the eight most significant pathogen-disease pairs. Each triangle is marked with a central dot, the color of which indicates whether the pair is a Tier 1 (gold) or Tier 2 (cyan) positive control, Expected Negative (black) control, or an unknown relationship (green).

Outside of our Tier 1 positive controls, the largest odds ratio (OR) obtained in the UKB is for systemic lupus erythematosus (SLE) with EBV (UKB OR = 3.98), which also has a large TNX odds ratio of 4.96. We also replicated the well-established connection between EBV and multiple sclerosis (UKB OR = 2.55; TNX OR = 4.45). Overall, HCV and HBV have the most replicated associations (25) of the pathogens tested (**Figure S4**). EBV, HBV, and HCV all have replicated associations in over ten different ICD10 blocks, reflecting the often systemic effects of infections by these pathogens.

Cytomegalovirus (CMV) has the most associations with an OR indicating risk (21 of its 24 replication associations have an OR greater than 1). In contrast, *Chlamydia trachomatis* has the largest number of protective relationships (15 of its 20 with an OR less than 1). “Other diseases of the urinary system” (ICD10: N39) had the most replicated associations across all pathogens (10), all of which are predicted to increase the risk of disease. The 3-character ICD10 code N39 includes both the communicable diagnosis “urinary tract infection” (N39.0) as well as several non-communicable forms of incontinence (N39.3, N39.4).

To characterize the replicated pathogen-disease pairs more broadly, we examined our results at the ICD10 block level, normally representing distinct body systems. *H. pylori* has the most replicated associations in a particular block: block 11 (K00 - K95), which contains diseases of the digestive system (**Figure S5**). In addition to the three Tier 2 positive controls discussed above, *H. pylori* is predicted to increase the risk of four additional digestive system diseases and to decrease the risk of “irritable bowel syndrome”, “diaphragmatic hernia”, “other diseases of esophagus”, and “gastro- esophageal reflux disease” (GERD).

### Phecode analysis of pathogen-disease associations

Phecodes are a phenotype encoding scheme developed originally for PheWAS studies^24,25^. Phecodes are sets of ICD10 codes bundled together under a single phenotype. They also include the use of exclusion criteria, which helps to reduce the presence of cases in a control cohort. To demonstrate the robustness of our pathogen-disease associations, we next repeated our ICD10-based analysis using Phecodes as the outcome variable instead of individual ICD10 codes. This analysis included 18,820 pathogen-Phecode unknown pairs, 10 Tier 1 pairs, and 104 Expected Negatives.

We first examined the positive control results, finding that 8 of the 10 Tier 1 results are significant in the discovery cohort (UKB data), all of which are also significant in the replication cohort (TNX data). As expected, this fraction of pairs that replicated pairs was much higher than the fraction observed for the Expected Negatives, 6 of the 104 pairs (**Figure 2b**). These results demonstrate that our Phecode-based model also has the discriminatory capacity to separate positive controls from negative controls.

After assessing the model performance, we next analyzed the unknown pathogen- Phecode pairs. Of the 18,820 unknown pairs, 3,289 (589 unique Phecodes) were significant in the discovery cohort. We were powered to test 2,784 of these pairs for replication in the TriNetX data, where we found that 1,341 pairs (449 unique Phecodes) fully replicated (**Supplemental Dataset 3**). Nearly all of the pairs with a Phecode corresponding to an ICD10 in Table 1 replicate, including multiple sclerosis with EBV, systemic lupus erythematosus with EBV, and ulcerative colitis with CMV, indicating agreement between the ICD10 results and the Phecode results.

### Orthogonal validations of the cytomegalovirus – ulcerative colitis relationship

We next sought orthogonal evidence supporting the 206 replicated associations identified by our approach. Virus-disease relationships are often reflected by higher expression levels of virus-encoded genes in patients compared to controls^26–30^.

Likewise, many viruses manipulate host gene expression patterns^31,32^, and the molecular processes of most complex diseases are now appreciated to be impacted by alterations to human gene expression levels^33,34^. We thus hypothesized that causative pathogen-disease relationships would be reflected in publicly available gene expression data. To test this hypothesis, we performed two complementary analyses. First, we examined viral gene expression levels in patients compared to controls. Second, we asked if genome-wide association study (GWAS) risk loci were enriched near human genes with altered expression levels following viral infection.

As a positive control, we first examined the well-established link between EBV and SLE. To this end, we identified six publicly available SLE case/control RNA-seq data sets performed in blood and B cell subsets (**Supplemental Dataset 4**). Collectively, these data contain 378 SLE cases and 74 control subjects. We used the VIRTUS software package^35^ to identify and quantify viral read counts in these data. As expected, this analysis revealed significantly higher EBV transcript levels in SLE cases compared to controls (p-value = 4.9E^-03^) (**Figure 4a**).

**Figure 4.**
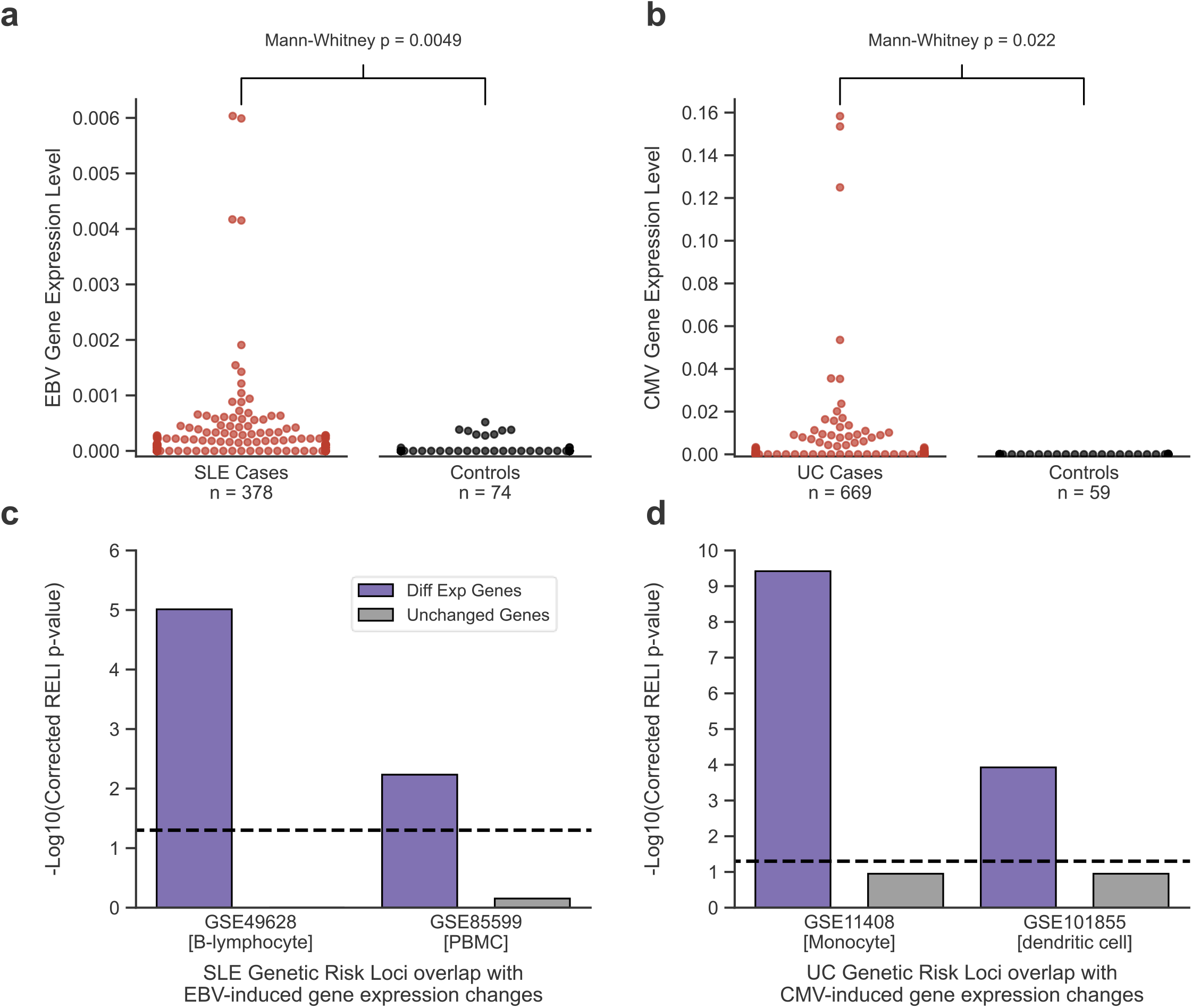
| Orthogonal validation of the EBV/SLE and CMV/UC associations. Orthogonal validation of the EBV/SLE (positive control) and CMV/UC (new prediction) associations using virus gene expression levels (top) and enrichment of disease risk loci near virus-induced differentially expressed human genes (bottom). **a** and **b**. Swarm plots showing viral read counts normalized by the size of the viral genome and the total number of human mapped reads in that sample, providing the final ‘Normalized Hit Rate’ calculated by the VIRTUS software package. Normalized hit rates were compared between cases and controls using a Mann-Whitney test with the p-values annotated on the plots: SLE versus controls (panel **a**) and UC versus controls (panel **b**). **c** and **d**. Bar plots indicating the enrichment of SLE (left, **c**) or UC (right, **d**) genome-wide association study (GWAS) loci proximal to genes with altered expression (purple bars) or unaltered expression (grey bars) after infection by the viruses EBV and CMV, respectively. Gene Expression Omnibus (GEO) ID and cell type are provided below each plot. The black dashed line indicates statistical significance (p = 0.05)

We next considered ulcerative colitis (UC), a disease with a suggestive but still unclear role for viral infection^36^. Our main analyses implicated both HIV and CMV in UC disease processes, with the replication cohort showing infection occurred before disease diagnosis. We thus examined RNA-seq data for a set of 669 UC cases and 59 controls obtained from seven studies using intestinal biopsies (**Supplemental Dataset 4**).

Similar to EBV and SLE, we observe significantly higher levels of CMV transcripts relative to controls in these samples (p-value = 2.2E^-02^) (**Figure 4b**), providing additional evidence of a role for CMV in UC disease processes.

As a second orthogonal analysis, we asked if GWAS disease risk loci are enriched near human genes with virus-induced expression level changes. To this end, we used our RELI algorithm^11^ to relate GWAS risk loci and public RNA-seq experiments examining virally infected and uninfected cells. In brief, this procedure uses a permutation-based method to estimate the significance of the overlap between the genomic coordinates of GWAS-identified risk loci and 200 kilobase windows around the transcription start site for genes with virus-altered gene expression levels (see Methods).

As a positive control, we first examined EBV and SLE. As expected, our analyses revealed significant overlap between SLE risk loci and genes that are differentially expressed upon EBV infection, in two independent studies performed in B lymphocytes and peripheral blood mononuclear cells (**Figure 4c**, purple bars; **Supplemental Dataset 5**). In contrast, when considering genes that did not change significantly upon infection (**Figure 4d**, grey bars), SLE risk loci are not enriched. These results are consistent with our previous observation that the genomic binding events of the EBNA2 regulatory protein, encoded by EBV, coincide with approximately half of all SLE risk loci^11^. Encouraged by these results, we next compared CMV-altered genes to UC genetic risk loci. Similar to the EBV-SLE results, we again observe highly significant overlap between CMV-altered genes and UC risk loci and insignificant overlap for expressed but unchanged genes (**Figure 4d**, purple and grey bars, respectively; **Supplemental Dataset 5**) in two different cell types (monocytes and dendritic cells). Similar results were obtained for the highly related Crohn’s disease and inflammatory bowel disease (**Supplemental Dataset 5**).

Collectively, these analyses provide compelling orthogonal evidence that the CMV-UC connection identified in both of our independent cohorts might represent a causative relationship.

## Discussion

In this study, we sought a broader understanding of the role played by pathogens in what are traditionally considered non-communicable diseases (NCDs). To this end, we developed a logistic regression model and applied the model to two large, independent biobank resources. Our model showed strong discriminatory performance on a set of positive and negative controls and replicated many additional well-documented pathogen-NCD associations. Overall, our results were robust regardless of the choice of International Classification of Diseases 10^th^ revision (ICD10) codes or Phecodes. We report evidence for over 200 new or previously tenuous pathogen-disease connections, including a role for cytomegalovirus (CMV) in ulcerative colitis (UC), which was supported by two orthogonal genomics-based validations, and provide the corresponding data on a freely accessible and easily browsable web server, https://tf.cchmc.org/pathogen-disease.

The relationship between *H. pylori* and gastroesophageal reflux disease (GERD) continues to be debated, with one recent meta-analysis reporting that the eradication of *H. pylori* increases the risk of GERD, thus indicating a possible protective effect^37^ and another recent systematic review rated the “evidence grade” for the association between *H. pylori* and GERD as low^37,38^. Our replicated result indicating that *H. pylori* has a protective effect adds additional evidence of this debated association. Outside of ICD10 block 11, *H. pylori* also has a risk relationship with “iron-deficiency anemia”, an association that has been published previously^39^, and a protective relationship with asthma, which has also been previously reported^40,41^.

Our results implicated both human immunodeficiency virus (HIV) and CMV in ulcerative colitis (UC). HIV has recently been linked to UC^42^. For CMV, a possible role is much less clear, perhaps due to the historical difficulty of differentiating between CMV colitis and inflammatory bowel diseases such as UC^36^. Although the hypothesis that CMV may be causal of UC remains contentious^43^, the ability of CMV to cause UC flare-ups is still heavily debated^44^. Our results should aid in these ongoing debates, including the replicated associations based on serologic and diagnostic data as well as our two orthogonal analyses, which all suggest a role for CMV in UC processes.

A recent study by Levine et al. took a similar approach to ours, with a specific focus on six neurodegenerative diseases^45^. Herein, we report results from an analysis across the disease spectrum. In addition to the comparatively limited scope of the Levine *et al.* study, looking at just six neurodegenerative diseases, there are several additional key differences between the 2023 Levine *et al.* study and ours. The Levine *et al.* study used hospital diagnosis codes as a pathogen proxy, some of which link to multiple pathogens, such as viral encephalitis. In comparison, we were able to pinpoint specific pathogens due to our use of serology data. Further, by restricting to inpatient hospital databases, the Levine *et al.* study focused on patients with infections sufficiently severe enough to require hospitalization. In contrast, our analyses included systematically measured titers for select UK Biobank (UKB) participants^46^, along with data from TriNetX (TNX), which pulls all available clinical laboratory test results for each patient, encompassing standard preventative screenings, outpatient diagnostic workups, as well as panels ordered during hospital stays. This is likely one reason why the odds ratios we report are much more modest than those reported by Levine *et al.* Thus, although both studies are of great utility, the results of the two studies are not directly comparable.

Roughly 40% (81/206) of the replicated associations in our study have odds ratios of less than one, indicating a potentially protective pathogen-disease relationship. For example, while high-risk strains of human papillomavirus (HPV) such as 16 and 18 are known to cause over 70% of cervical cancer, our results also suggest that HPV-16 and - 18 can be “protective” for diseases such as “seborrheic dermatitis” and “other dermatitis”. Indeed, viral infections that increase the risk of one phenotype or disease can reduce the risk for others^47–50^. More generally, the role of viral infection in shaping the human immune system and subsequent immune responses has been studied extensively. It is well appreciated that viral infection can rewire the chromatin of immune cells and shape subsequent responses of a person toward additional inflammatory insults^51,52^. For example, a viral response that results in an interferon-based immune response could be protective in the context of diseases driven by T cell helper-2 type inflammation^53^. Vaccination against a particular pathogen will protect against infection and, thus, the disease risks associated with that pathogen. However, further studies will be required to investigate whether vaccination will confer the same protective effects against NCDs we report in this study.

The availability of the large datasets from the UK Biobank and TriNetX enabled this research. The associations identified in our study depend upon a sufficient number of subjects that have both accompanying diagnostic and serology data. As additional larger datasets are released, it will be critical to validate this study’s findings and use the additional statistical power to examine NCDs for which we were not powered. Identifying potentially causal etiological mechanisms driving these pathogen-disease associations will also be important, as recently exemplified by the Epstein-Barr virus(EBV) – multiple sclerosis (MS) field^11,12,51,54,55^. Furthermore, the goal of this study was to attempt to identify connections between pathogens and non-communicable diseases.

Combinations of pathogens can also have impacts on human disease etiology. For example, Plasmodium falciparum and EBV have been shown to increase the risk of endemic Burkitt lymphoma synergistically^56–58^. Likewise, genetics likely plays a vital role in pathogen-host interactions. Future studies applying new methodologies to much larger cohorts than those presented here will be important for identifying novel combinations of pathogens and host genetic variant-pathogen interactions that impact human disease.

Although attempts were made to minimize limitations in this study, some remain. For example, by their very nature, electronic health records and biobank data are noisy. We attempted to address this noise by examining two independent datasets covering vastly separate geographic locations, requiring a disease-pathogen pair to be significant in both, to be considered significant. Another limitation is that the datasets used in this study are based on cohorts from only two countries (the United States and the United Kingdom). Accordingly, the 20 pathogens investigated are largely prevalent and of most importance to the people of those regions as compared to the rest of the global population. An additional limitation of the study includes the possibility of cross-reactivity occurring while testing for a particular pathogen. While each of the serological tests was approved by the Clinical Laboratory Improvement Amendments program and used to inform clinical care of patients, it is possible that some non-viral human protein epitopes cross-reacted with the viral antigens in the serological tests^12,54,59^. Finally, not all confounders in the UK Biobank models could be adjusted for in the replication cohort because the data were unavailable.

In summary, we present the largest systematic assessment to date of pathogens in the context of non-communicable human disease. Using complementary discovery and replication datasets, we identified 206 replicated pathogen-disease relationships, including additional orthogonal evidence strongly supporting a relationship between CMV infection and ulcerative colitis. We anticipate that this rich data resource will form the foundation for future characterization of the many currently unknown pathogen-disease relationships.

## Methods

### UK BioBank cohort

The UK Biobank (UKB) is a prospective cohort study containing medical, sociodemographic, and genetic data for nearly 500,000 adults from across the United Kingdom^60^. In this study, diagnoses for all participants were extracted from two UKB fields, “First Occurrences” [UKB Category 1712], which is a synthetic field generated by UKB analysts that collates diagnoses from primary care, hospital inpatient, death registry, and self-reported records, and “Type of Cancer – ICD10” [UKB Field 40006], which contains cancer diagnoses extracted from national cancer registries for linked participants. The first occurrences data are limited to 3-character International Classification of Diseases 10^th^ revision (ICD10) codes, e.g., M32.9 is recorded as M32, so the cancer registry diagnoses were truncated to match. Analyses were limited to diseases with at least 17 cases and 187 total samples within the cohort (see power calculation below).

Data for 45 antibody titer levels representing immune responses to 20 unique pathogens [UKB Category 1307] were downloaded and Log10 transformed before analysis. Antibody titer measurements were performed using a multiplex serology approach based on the enzyme-linked immunosorbent assay (ELISA) concept as described by Mentzer *et al.*^46^. Similar to Mentzer *et al.,* a series of 10 additional health-related and sociodemographic variables that could potentially be associated with both disease status and antibody titer level were collected to test for confounding during analyses. The continuous covariates age and body mass index (BMI) were scaled by a factor of 10, while all other covariates were either already categorical or were discretized (**Table S1)**. The scikit-learn IterativeImputer method, based on the Multivariate Imputation by Chained Equations (MICE) algorithm, was used to impute missing covariate values. Thirty-two participants were missing BMI values and eight were missing Townsend deprivation index values.

### TriNetX cohort

TriNetX, LLC (TNX) is a private organization that has built a global medical research network that enables healthcare organizations to make their electronic health record (EHR) data more easily accessible to researchers in a de-identified manner, enabling Real World Data analyses. To prevent possible outlier values due to results being reported in different units or encoding errors, continuous laboratory test results were excluded, thereby limiting our analysis to only binary tests where the results were either positive or negative. A query built by combining a complete list of Logical Observation Identifiers Names and Codes (LOINC) codes that corresponded to binary clinical laboratory tests for our pathogens of interest was run on 02-14-2023 across 73 healthcare organizations on the TNX Research Network. The search resulted in a list of just over 11 million unique patients who had in their EHR a result for at least one of the laboratory tests in our query. Diagnoses for all participants were collected, and cohorts were automatically generated for each pathogen-disease pair. The use of GNU Parallel^61^ made the processing of the immense amount of TNX data much more tractable. For a specific pathogen-disease pair, those with a result for a particular LOINC code but without the diagnosis of interest were considered controls, and only those with a test result (positive or negative) before the earliest diagnosis for the diagnosis of interest in their medical record were considered cases. We removed those with the diagnosis appearing in the EHR before the laboratory test, as the temporal relationship of infection prior to disease diagnosis could not be firmly established for them. We attempted to extract all potential confounding variables considered in the UKB analysis from the TNX data; however, data for only three of the ten were available. For situations where a covariate was included in the UKB model but was not available in the TNX data, the covariates were dropped from the TNX model before refitting. After finding few ICD10 B24 cases in TNX (a diagnosis used in UKB to indicate human immunodeficiency virus (HIV) infection, “Unspecified human immunodeficiency virus [HIV] disease”), it was determined that in the United States (the primary source of TNX data) the ICD10 code B20 appears to be primarily used to indicate HIV infection. Thus, the TNX results for B20 were merged with the UKB B24 results. However, this is the only ICD10 code for which this was done.

### Phecode analysis

Phecodes were generated with the PheWAS R library (v0.99.6.1; R v4.0.2) and a minimum code count of one using previously published methods^62–64^. The resulting Phecodes were filtered to remove those with a disease group of infectious diseases, injuries & poisonings, congenital anomalies, or symptoms. We applied the same statistical analysis to the Phecodes that we used for ICD10 codes, including the same study design involving both the UKB and TNX cohorts, requiring the same statistical thresholds to be met (UKB per-Phecode false discovery rate (FDR) < 0.3 and TNX per-Phecode FDR < 0.01). Due to technical issues, we were unable to include the requirement for a laboratory result to be present in the EHR before diagnosis in the TNX cohort. A set of infectious disease Phecodes were used as positive controls (Tier 1 only) or negative controls (Expected Negatives) – see Supplemental Dataset 3.

### Establishment of the minimum number of cases and total samples for analysis

To ensure that the comparisons made had sufficient statistical power to identify associations if present and minimize the multiple testing correction burden, we sought to determine what a minimum number of cases and controls would be. While our analytical strategy used a logistic regression model, to determine a preliminary power estimate, we opted to consider the power to detect differences in antibody levels between those with and without disease for known antibody-disease pairs. This approach enabled us to calculate the effect sizes (mean difference between groups divided by pooled standard deviation) and evaluate the power required to detect such differences. While the logistic regression models (especially with covariates) likely have different power estimates, our goal was to identify a pragmatic threshold for the minimum number of cases with a reasonable likelihood of success. We looked at effect sizes across 14 known positive antibody-disease pairs in the UKB data, which collapsed to eight pathogen-disease pairs. These pairs constitute our “Tier 1” positive controls and represent those infectious diseases that are directly caused by a pathogen, such as “herpes zoster (HZ)” (ICD10: B02) diagnosis and varicella-zoster virus, the virus that causes HZ. Effect sizes at the antibody-disease level ranged from 0.1 to 3.9. After collapsing the antibody results to the largest effect per pathogen-disease pair, effect sizes ranged from 0.2 to 3.9, with a median effect size of 0.72. Using the G*Power 3.1 software package^65^, we calculated the number of cases required for 80% power with an alpha of 0.05, assuming ten controls per case, yielding a minimum case number of 17.

We recognize that more controls may be available, but higher numbers of controls impact power only slightly (data not shown). Further, using the median effect size of known positives might underestimate the power for some pathogen-disease associations; this conservative choice was made to balance the inclusion of diseases while ensuring sufficient statistical power.

### Model development and application

We modeled the association between a given disease and pathogen using a logistic regression model with disease status as the binary outcome and the pathogen proxy value (continuous for UKB antibody titers, categorical for TNX positive/negative binary lab tests) as the predictor (**Formula 1**). Since each cohort uses a different type of predictor, we note that the odds ratio (OR) has a slightly different meaning between the two. In the UKB cohort, the OR represents the increase in odds of developing a disease per 10-fold increase in antibody titer level. Whereas in the TNX cohort, the OR represents the increase in odds of developing a particular disease in those subjects after infection by a given pathogen. All sex-specific diseases had controls limited to those participants at risk, e.g., cervical cancer (ICD10: C53) used only females without a cervical cancer diagnosis as controls. For all pregnancy and childbirth-related diseases, only patients with a record of a healthy birth (ICD10 codes O80 - O84) were used as controls (i.e., patients with records of both a healthy birth and the diagnosis of interest were removed from the analysis to prevent them being used as both cases and controls).

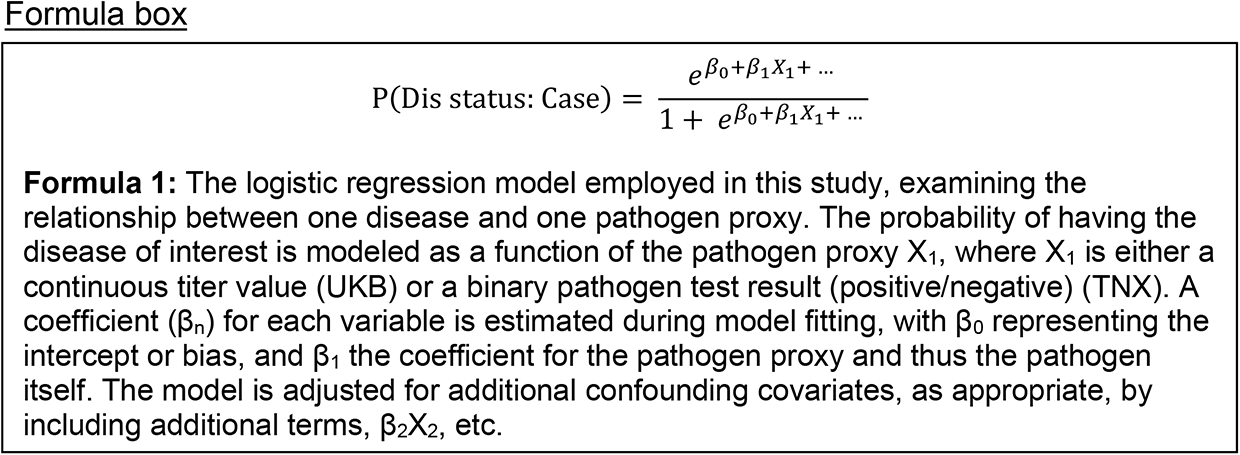

After adjusting our logistic regression model for all covariates found to be significantly associated with both disease status and pathogen proxy in separate univariate statistical tests (**Table S2**), we ran a backward elimination procedure (stepAIC, MASS R library), removing any non-significant covariates from the final model, to mitigate possible overfitting. We attempted to replicate models that showed statistical significance in the UKB cohort using the replication cohort by simply refitting the same model with the TNX data.

In the rare situation where a categorical pathogen test (TNX) had five or fewer patients in one of the cells of the pathogen-disease contingency table, we altered our model slightly. For example, for the diagnosis of “herpesviral [herpes simplex] infections” (ICD10: B00) and the pathogen test with LOINC code 93439-8, there is only one disease control with a positive test result for herpes simplex virus 1, the cause of this disease. This represents a strong association that would be lost with a standard logistic regression model. Thus, in these situations, we instead employed Firth’s bias-reduced logistic regression method^66^ (R library logistf), which is appropriate in such situations.

A permutation procedure was performed to verify the robustness of the UKB model results. Briefly, 10,000 permutations were run for each antibody-disease pair (45 total pairs per disease), where disease status was randomly shuffled amongst the participants while keeping the number of cases and controls and the model itself, i.e., confounders adjusted for, constant. Next, all permutation results for a particular disease were pooled into a larger per-disease null distribution now containing 450,000 permutation results. Empirical p-values for each antibody-disease pair were calculated by comparing the nominal p-value from our analytical model to the respective per-disease null distribution.

We applied a per-disease Benjamini-Hochberg (BH) false discovery rate^67^ at the pathogen-disease level to control for multiple testing. Since we were using a discovery-replication model, we used a lenient FDR threshold of 0.3 for our discovery cohort to reduce the likelihood of false negatives that might occur due to the smaller size of this cohort. Despite the smaller number of subjects, the UKB cohort has the advantage of systematic measurements of antibody titers. We therefore used a “lenient” FDR threshold in the UKB cohort to minimize initial false negatives, applying a much more stringent FDR threshold of 0.01 to our larger replication cohort to minimize false positives. Our Tier 1 analysis drove the choice of a precise 0.3 cutoff in our discovery cohort. All eight Tier 1 associations were significant in the UKB data at this cutoff, which is expected given their well-accepted causal relationships. We emphasize that many Tier 2 associations identified by a semi-automated literature search might be incorrect or might represent weaker associations that are harder to detect. We therefore did not require all Tier 2 associations to be significant in the UKB data and instead expected an intermediate replication rate for the Tier 2’s between the Tier 1 and Expected Negatives.

### Model assessment

To assess the model’s performance, we calculated associations for a set of positive and negative control pairs. We used the Tier 1 controls as described above for the positive controls. Six infectious diseases are included in the Tier 1 controls, two of which can be caused by two different pathogens included in this study (“herpesviral [herpes simplex] infections” by herpes simplex virus 1 or 2 and “unspecified viral hepatitis” by hepatitis B (HBV) or hepatitis C (HCV)). The negative control set, termed “Expected Negatives”, represents the complement of the Tier 1 controls, e.g., a herpes zoster diagnosis paired with HBV instead of the causal agent, varicella zoster virus (VZV). As an additional assessment, a second set of positive controls using only non-communicable diseases (NCDs) (“Tier 2”) was collected using a semi-automated literature mining approach. In brief, we employed the log product frequency (LPF), a previously published method for quantifying the co-occurrence of search terms in PubMed^68,69^. Specifically, we employ a negated form of LPF to rank pathogen-disease pairs by the number of PubMed co-citations (disease and pathogen), normalized by the number of citations of each separately (**Formula 2**). The negation rotates the LPF values around the origin, allowing us to deal with LPF values greater than zero, whereas regular LPF values are all less than or equal to zero. The closer the LPF value is to zero, the more the disease and pathogen were co-cited as opposed to cited individually.

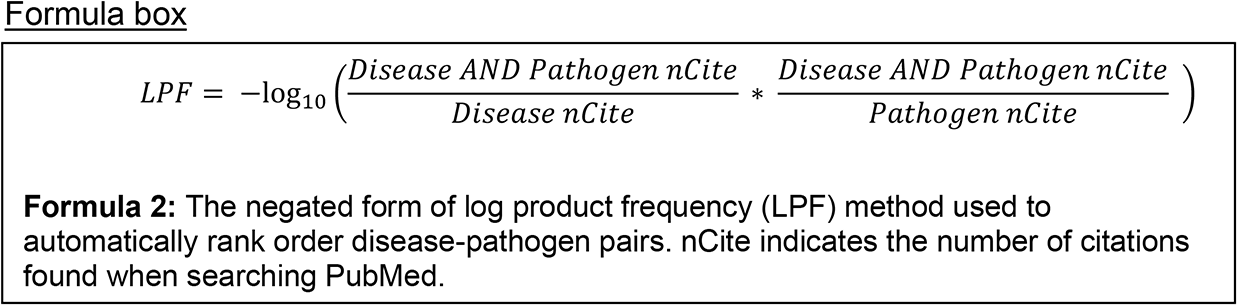

Searches of PubMed were conducted using the Entrez functions from the Python library Biopython using default settings on 8/11/2020. The top 175 results ranked by LPF (**Supplemental Dataset 6**) were manually reviewed by M.L. The evidence column in Supplemental Dataset 6 contains one of four levels used to gauge the literature support found for a particular pathogen-disease pair during the manual review. Briefly, a rating of “High” indicates that at least one published meta-analysis supporting the association was found, or the connection is textbook knowledge (e.g., Hepatitis B causes hepatitis). “Medium” signifies that some cross-sectional or longitudinal studies were found supporting the pair’s association. An evidence level of “Low” indicates that only a handful of case reports pointing to a possible association were found. “None” means no papers were found indicating an association.

After limiting pairs to only those with a “high” evidence grade, we were left with 83 pathogen-disease pairs, making up our “Tier 2” positive controls. Pairs with less evidence and thus not rated as “high” were classified as either “medium”, “low”, or “none”, for several reasons, all of which were excluded from the Tier 2 list. For example, D50, “iron deficiency anemia” paired with Merkel cell polyomavirus (MCV), ranked 22^nd^ by LPF, was marked as “none” after manual review. Notably, the acronym MCV can also stand for “mean corpuscular volume”, a blood measurement that can indicate iron deficiency anemia, which is likely the cause for the high ranking. Further, K58, “irritable bowel syndrome (IBS)” paired with *H. pylori*, ranked 138^th^ by LPF, was classified as having low evidence by M.L. after the manual literature review. Although many studies have examined this association, the two most recent, i.e., published before our literature search, meta-analyses^70,71^ have found no significant association between the two. Further, since the initial literature review, two additional meta-analyses found non-significant associations between IBS and *H. pylori* as well^72,73^.

This semi-automated approach was required because a manual literature review for (426 diseases x 20 pathogens = 8,520) disease-pathogen pairs was intractable. The raw automated queries and results, including the number of citations and PubMed ID (PMID) lists for pathogen-disease co-citations, pathogen only, and disease only, can be found in **Supplemental Dataset 7**.

### Orthogonal validation of CMV-UC association

We attempted to validate two of our replicated findings using two distinct ‘omics-based methods.

First, to capture differences in viral gene expression levels between cases and controls for the diseases of interest, we used publicly available RNA-seq data from case/control cohorts and the bioinformatics tool VIRTUS v1.2.1^35^ using default parameters except reducing the default “hit_cutoff” from 400 to 0 since we were only investigating a small number of pathogens. Briefly, VIRTUS is a pipeline that takes an input RNA-seq FASTQ file and aligns the reads to a reference human genome (hg38; GCF_000001405.39) using STAR^74^. It then attempts to align any unaligned reads to a second pre-compiled index file containing a user-specified set of viral genomes, here Epstein-Barr virus (EBV) (NC_007605.1) and cytomegalovirus (CMV) (NC_006273.2). The resulting number of mapped reads for each virus was first normalized by the pathogen’s genome length, then normalized by the number of mapped human reads, providing the flexibility to compare results across both different samples and experiments as well as different pathogens. Finally, the non-parametric Mann-Whitney U test was used to compare the normalized read counts for a particular pathogen between cases and controls.

Second, to examine if genome-wide association study (GWAS) loci for the diseases of interest were enriched near human genes that have altered expression levels upon infection by the pathogens of interest more so than unchanged genes, we used our RELI tool^11^. RELI estimates the statistical significance of the overlap between a set of input genomic loci and a peak file, typically GWAS loci and ChIP-seq peaks. It does this through a permutation-based procedure by first counting the number of overlaps between the input loci and peaks, then permuting the input loci around the genome and again counting the number of overlaps with the peaks, building a null distribution.

Finally, to calculate significance, RELI compares the total number of overlaps seen in the input loci set to the constructed null distribution.

We obtained GWAS data for ulcerative colitis (UC) and systemic lupus erythematosus (SLE) from the NHGRI-EBI GWAS catalog (v1.0.2-associations_e96_r2019-05-03)^75^. A genome-wide significance cutoff of 5x10^−8^ was used, and we considered only data from European populations due to the prevalence of GWAS data for this ancestry group.

Independent loci were identified for each phenotype using linkage disequilibrium (LD)- based pruning with PLINK^76^ (window size 300,000 kb, SNP shift size 100,000 kb, and r^2^ < 0.2). These independent loci were then expanded to incorporate variants in strong LD (r^2^ > 0.8) again using PLINK. Next, we downloaded lists of genes with expression changes in response to CMV or separately EBV infection from the VExD database (https://vexd.cchmc.org)^77^ (**Supplemental Dataset 8)**. Differentially expressed genes (DEGs) are identified in VExD as those genes with an adjusted p-value < 0.05 and absolute fold change > 2, when comparing infected and uninfected cells of the same type within a single study. As a null model, for each study, we also identified sets of genes that, while expressed, do not significantly change upon infection (adjusted p >= 0.05, and fold change < 1.2) and randomly selected the same number of genes to match the corresponding differentially expressed gene set. We then ran RELI using the GWAS risk loci for each disease as input against the genomic regions defined by a 200kb window centered on the transcription start site of each input gene.

## Data Availability

The data used for the main analysis from The UK Biobank can be accessed via an application for Tier 2 UKB data. Further the data from TriNetX (https://live.trinetx.com accessed on 14 February 2023) can be requested directly from TriNetX. In both cases costs may be incurred and a material transfer agreement or data sharing agreement is required. NCBI GEO GSE IDs and the lists of GWAS loci from the NHGRI-EBI GWAS Catalog used for orthogonal analyses can be found in the included supplementary datasets.

https://www.ukbiobank.ac.uk

https://trinetx.com

https://www.ebi.ac.uk/gwas

https://www.ncbi.nlm.nih.gov/geo

## Acknowledgments

We thank Ivan Marazzi, Chris Benner, Brad Rosenberg, Emily Miraldi, Juan Fuxman- Bass, and Artem Babaian for their insightful comments on the manuscript. We also thank Matthew Skowronek, Eric Young, Jeff Warnick, and the rest of the engineering team at TriNetX for additional assistance working with the TriNetX data. We thank Monika Grabowska, Wei-Qi Wei, and Spiros Denaxas for their invaluable help with the Phecode analysis. We thank The University of Cincinnati Center for Clinical and Translational Science and Training (CCTST) for assistance with accessing the TriNetX data.

This research was conducted using the UK Biobank Resource under application number 47377.

TriNetX, LLC is compliant with the Health Insurance Portability and Accountability Act (HIPAA), the US federal law which protects the privacy and security of healthcare data, and any additional data privacy regulations applicable to the contributing healthcare organization.

CCTST is supported by the National Center for Advancing Translational Sciences of the National Institutes of Health (NIH), under Award Number 2UL1TR001425-05A1. The content is solely the responsibility of the authors and does not necessarily represent the official views of the NIH.

This research was funded by National Institutes of Health (NIH) R01 DK107502, R01 AI148276, U01 HG011172, U19 AI070235, and P30 AR070549 to L.C.K.; R01 HG010730, R01 GM055479, and U01 AI130830, to M.T.W.; R01 AR073228, R01 NS099068, and R01 AI024717 to M.T.W. and L.C.K.; R01 CA226802 to N.S. Funding was also provided by Cincinnati Children’s Hospital Medical Center ARC Award 53632 to M.T.W. and L.C.K.

## Author Contributions

Conceptualization: M.L., L.C.K., M.T.W.; Study design: M.L., L.C.K., M.T.W.; Statistical analysis design: M.L., D.S., L.M., M.T.W.; Computational analysis: M.L., S.P.; Writing: M.L., L.C.K., M.T.W.; Review and approve final manuscript: M.L., D.S., S.P., N.S., B.H.,

L.M., L.C.K., M.T.W.; Funding: N.S., L.C.K., M.T.W.

## Code Availability

All code used in this project is available from the Weirauch Research Lab’s GitHub page, https://github.com/WeirauchLab/pathogen_ncd.

## Data Availability Statement

The data used for the main analysis from The UK Biobank (UKB) can be accessed via an application for Tier 2 UKB data. Further, the data from TriNetX (https://live.trinetx.com, accessed on 14 February 2023) can be requested directly from TriNetX. In both cases, costs may be incurred, and a material transfer agreement or data-sharing agreement is required. NCBI GEO GSE IDs and the lists of GWAS loci from the NHGRI-EBI GWAS Catalog used for orthogonal analyses can be found in the included supplementary datasets.

---

**Figure S1.**
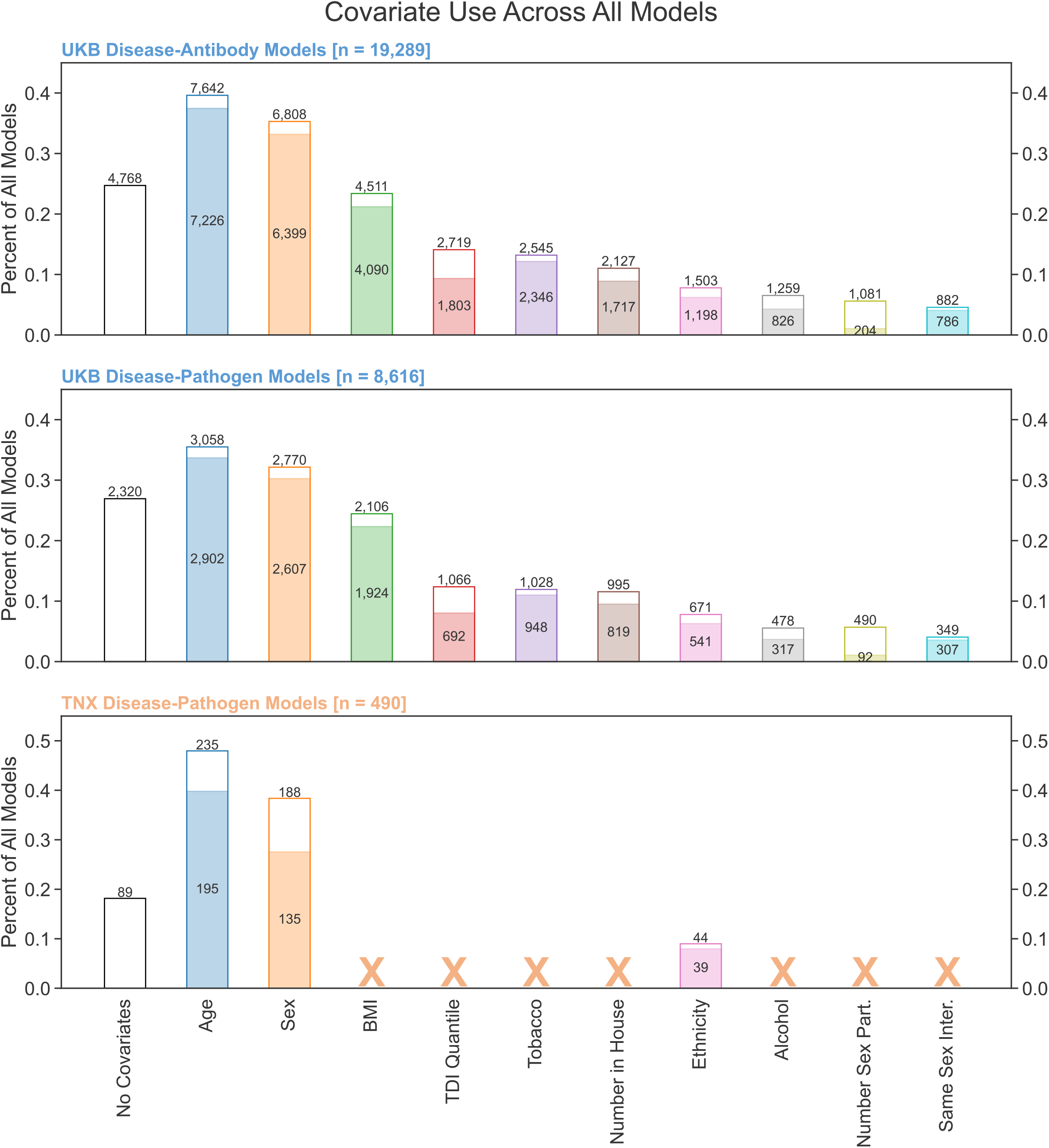
| Covariate use across models. Bar charts summarizing covariate usage in antibody-disease and pathogen-disease tests across the discovery and replication cohorts. The height of each bar indicates the percentage of all models that were adjusted for the corresponding sociodemographic or health-related covariate. To be included in an antibody-disease model for adjustment, a covariate had to be significantly associated (unadjusted p < 0.05) in univariate tests with both the disease status and the titer level separately (Table S2). The filled portion of each bar indicates the percent of the final multivariate logistic regression models in which the covariate remained significant (unadjusted p < 0.05). The covariates that were unavailable in the TriNetX (TNX) cohort are marked by an orange “X”. Abbreviations: Part: Partners; Inter: Intercourse; TDI: Townsend deprivation index.

**Figure S2.**
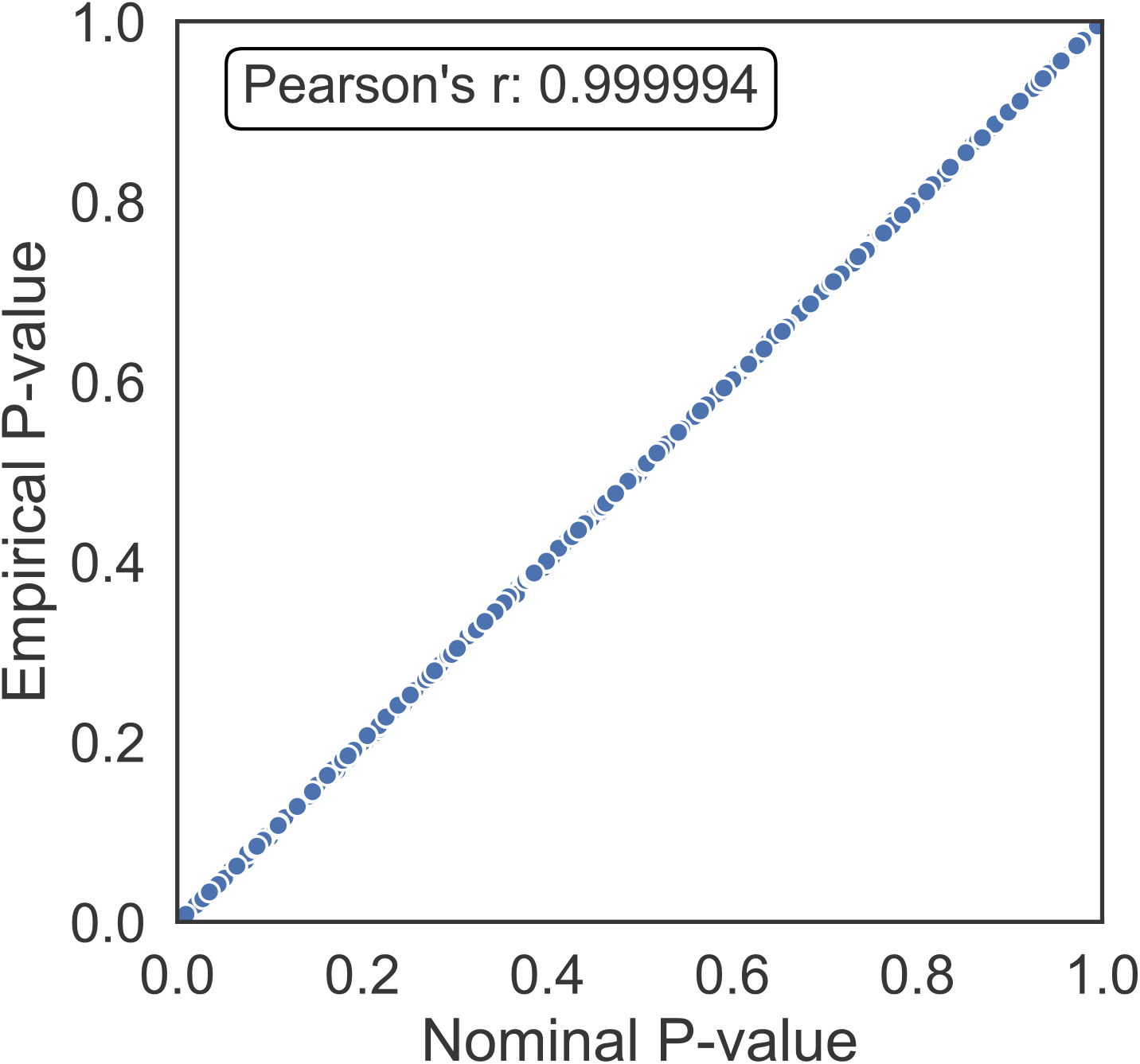
| Nominal versus empirical p-values across all UK Biobank antibody-disease models. Scatter plot comparing the nominal and empirical p-values for all discovery cohort antibody-disease models. To calculate empirical p-values, 10,000 permutations of each antibody-disease model were performed. All permutations for a particular disease were combined into a per-disease null distribution, yielding 450,000 permutation results per disease (see Methods). The nominal p-value for a specific antibody-disease model was compared to the disease-specific null distribution to calculate the empirical p-value.

**Figure S3.**
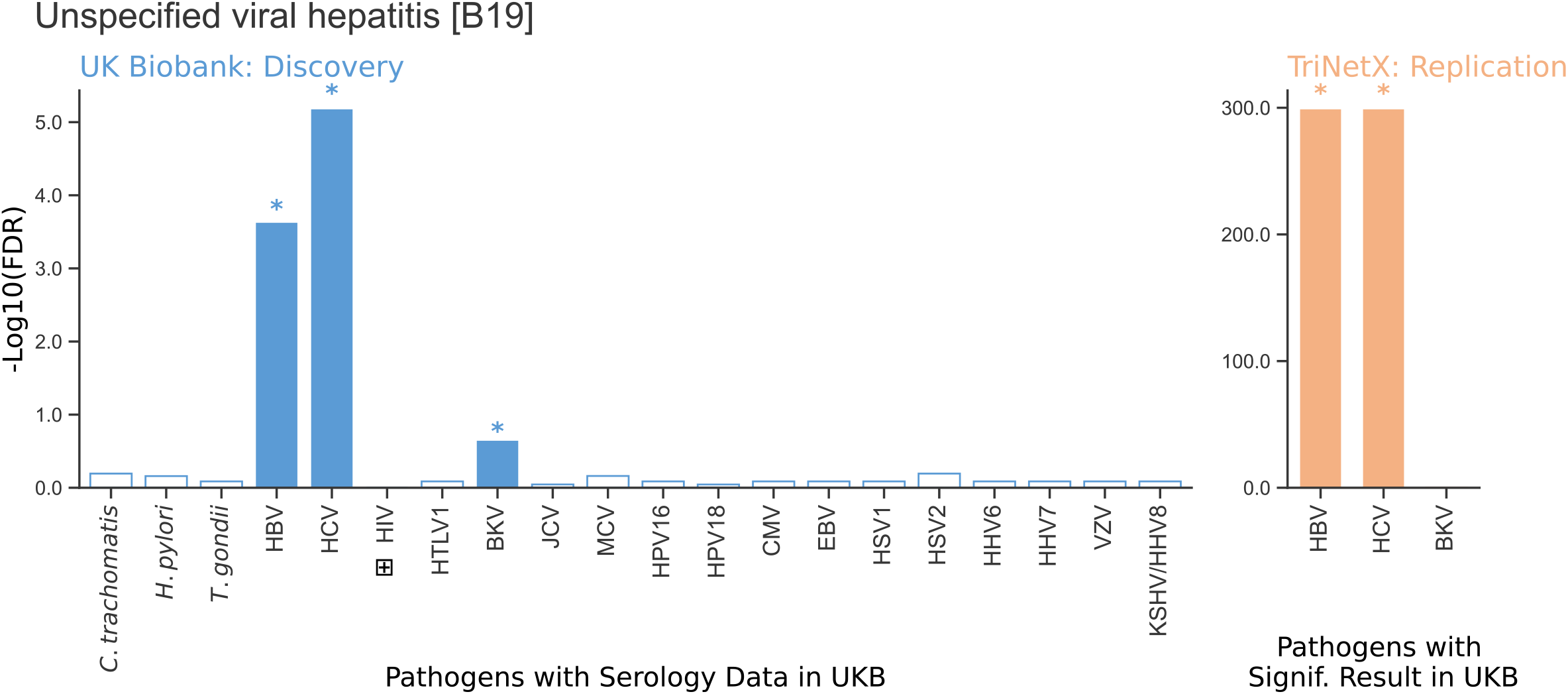
| Illustration of the two-step discovery-replication process. Bar charts depicting our methods’ results for a control disease. The left plot (blue) displays the significance of association (-Log10 transformed per-disease Benjamini- Hochberg false discovery rate (FDR)) for each pathogen with the disease of interest, here the control disease, “unspecified viral hepatitis”. Significant associations are depicted as filled bars with colored asterisks above. Hepatitis B (HBV), hepatitis C (HCV), and BK Virus (BKV) are all significantly associated with “unspecified viral hepatitis” in the UK Biobank (UKB) cohort. The right plot (orange) shows the results of testing only the significant UKB results in the replication cohort, TriNetX. Only HBV and HCV remain significant at the more stringent replication threshold of per-disease FDR < 0.01, leaving the pairs HBV-unspecified viral hepatitis and HCV-unspecified viral hepatitis the only replicated results.

**Figure S4.**
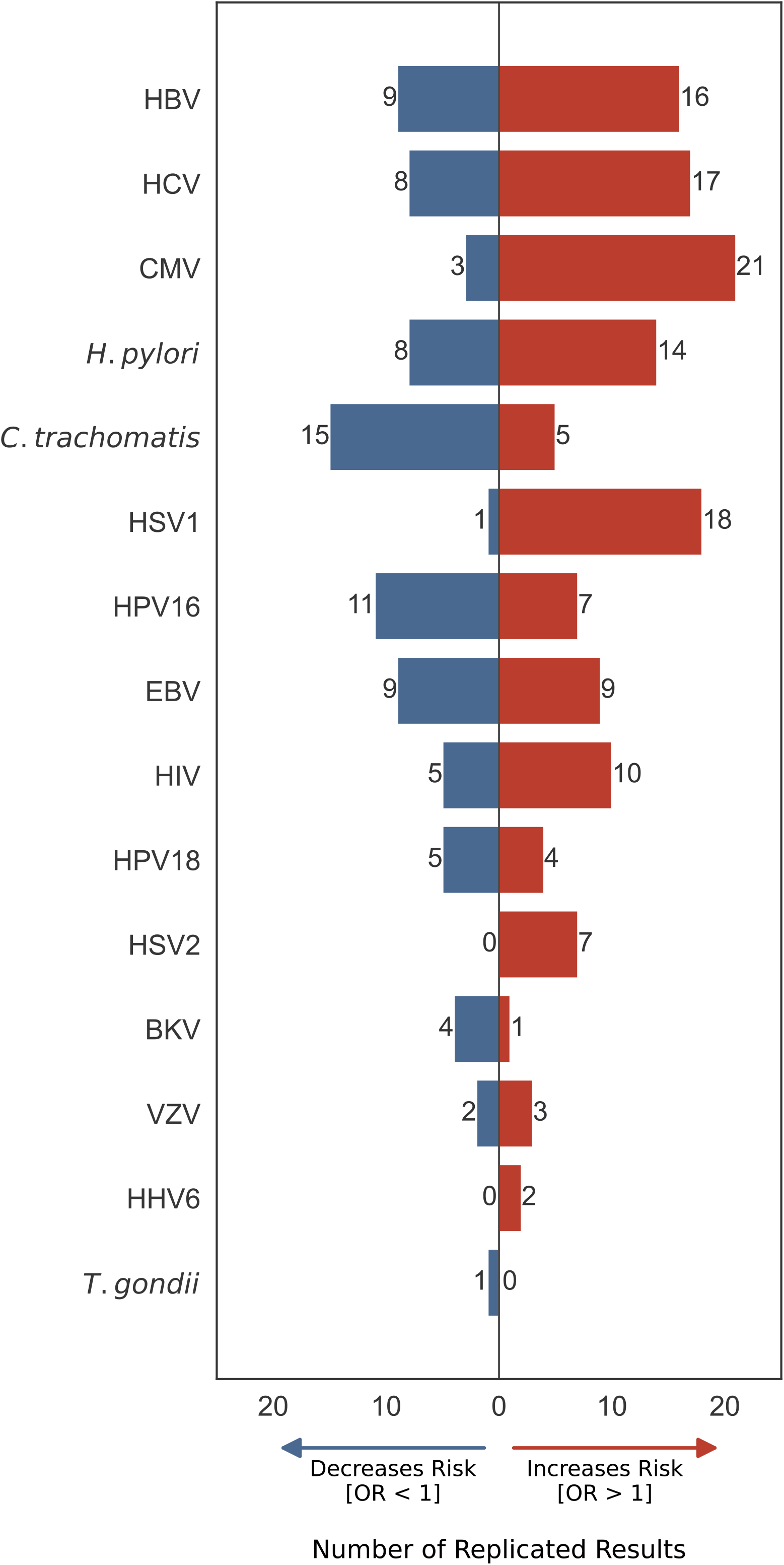
| Replicated results split by pathogen and effect direction. A split bar chart showing the number of replicated “risk” and “protective” associations for each pathogen. The total number of replicated results for each pathogen is split by effect direction. Those with a UK Biobank (UKB) and TriNetX (TNX) odds ratio of less than one are represented by the blue bars to the left of the center line. While those with a UKB and TNX odds ratio greater than one are represented by the red bars to the right of the center line.

**Figure S5.**
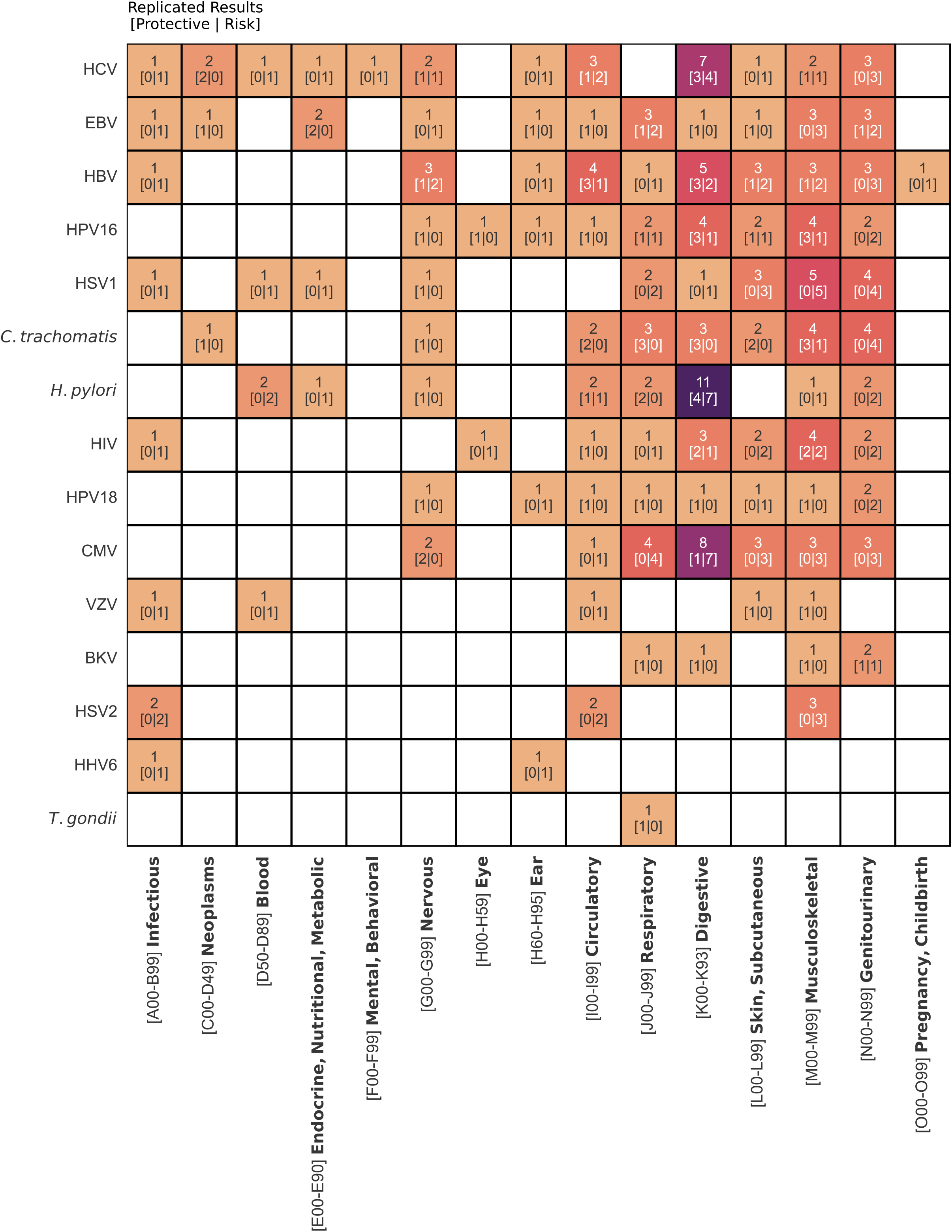
| Heatmap of replicated results at the ICD10 block level. A heatmap showing the total number of replicated results for each pathogen across all diseases in an International Classification of Diseases 10th revision (ICD10) block, which generally includes only diseases of a particular body system. All cells are annotated with the total number of replicated results above, in square brackets, the number of these replicated results with odds ratios less than one, followed by those with odds ratios greater than one. White squares with no annotation indicate that no replicated associations were found for that pathogen and any tested diseases in that ICD10 block.

**Table S1.** | Summary statistics for additional covariates across both cohorts. This table contains the summary statistics for the ten health-related and sociodemographic variables considered as possible confounders. The mean, standard deviation, and range are presented for continuous variables. For categorical variables, counts for each group are included. Note that the resolution for age differed between cohorts, with the UK Biobank having a resolution of one month, while TriNetX only provided the year of birth.

|  |  | UK Biobank | TriNetX |
| --- | --- | --- | --- |
| Sex | Female | 5,276 | 7,410,388 |
|  | Male | 4,153 | 3,807,258 |
|  | Other | 0 | 5,827 |
|  | Total | 9,429 | 11,223,473 |
| Age |  |  |  |
| Resolution: | Mean | 67.5 | 46.3 |
| UKB: Month | SD | 8.1 | 17.3 |
| TNX: Year | Range | [43.8 - 82.6] | [0 - 91] |
| Race/Ethnicity |  |  |  |
|  | White | 8,883 | 4,654,725 |
|  | Asian | 231 | 258,886 |
|  | Black | 140 | 1,664,492 |
|  | Hispanic | - | 860,830 |
|  | American Indian or Alaska Native | - | 36,516 |
|  | Native Hawaiian or Other Pacific Islander | - | 12,767 |
|  | Other | 175 | 3,735,257 |
|  | Total | 9,429 | 11,223,473 |
| BMI |  |  |  |
|  | Mean | 27.3 | - |
|  | SD | 4.8 | - |
|  | Range | [16.2 - 61.3] | - |
| n Imputed |  | 32 | - |
| Townsend Deprivation Index |  |  |  |
|  | Lowest 1/6 | 1,572 | - |
|  | Middle 2/3 | 6,285 | - |
|  | Highest 1/6 | 1,572 | - |
|  | Total | 9,429 | 0 |
| n Imputed |  | 8 | - |
| Number in House |  |  |  |
|  | Alone | 1,712 | - |
|  | 2 | 4,331 | - |
|  | 3 | 1,524 | - |
|  | 4 | 1,282 | - |
|  | >= 5 | 514 | - |
|  | Unknown | 66 | - |
|  | Total | 9,429 | 0 |
| **Tobacco Use** |  |  |  |
|  | Never | 5,210 | - |
|  | Previous | 3,218 | - |
|  | Current | 949 | - |
|  | Unknown | 52 | - |
|  | Total | 9,429 | 0 |
| **Alcohol Use** |  |  |  |
|  | Never | 417 | - |
|  | Previous | 332 | - |
|  | Current | 8,661 | - |
|  | Unknown | 19 | - |
|  | Total | 9,429 |  |
| **Number of Sex Partners** |  |  |  |
|  | 0 partners | 88 | - |
|  | 1 partner | 2,265 | - |
|  | 2-3 | 1,838 | - |
|  | 4-5 | 1,255 | - |
|  | > 5 | 2,343 | - |
|  | Unknown | 1,640 | - |
|  | Total | 9,429 | 0 |
| **Same-sex Intercourse** |  |  |  |
|  | No | 8,204 | - |
|  | Yes | 283 | - |
|  | Unknown | 942 | - |
|  | Total | 9,429 | 0 |

**Table S2.** | Univariate tests used to determine confounding. This table shows the type of univariate test performed between each covariate and antibody titer and each covariate and disease status to determine if the covariate was confounding. For a covariate to be considered a confounder, it had to be significantly associated (unadjusted p < 0.05) separately with both disease status and antibody titer.

| Covariate | Ab Titer Assoc Test | Disease Status Assoc Test |
| --- | --- | --- |
| Sex | t-test | Chi-squared |
| Age | linear regression | t-test |
| BMI | linear regression | t-test |
| Race | ANOVA | Chi-squared |
| Townsend Deprivation Index | ANOVA | Chi-squared |
| Number in House | ANOVA | Chi-squared |
| Tobacco Use | ANOVA | Chi-squared |
| Alcohol Use | ANOVA | Chi-squared |
| Number of Sex Partners | ANOVA | Chi-squared |
| Same-sex Intercourse | ANOVA | Chi-squared |

## Supplemental Datasets

Supplemental Dataset 1 | Overview of diseases outside of ICD10 chapter 1 with an infectious component.

All diseases were tested at the 3-character International Classification of Diseases 10^th^ revision (ICD10) code level. This document contains a review of whether those diseases outside of ICD10 chapter 1 (“infectious diseases”) are either infectious at the 3-character level, at a sub-code level, or not infectious at all. For example, J11, “influenza, virus not identified” is infectious at the 3-character level, whereas N39, “other disorders of urinary system” is not infectious at the 3-character level but does include the sub-code N39.0, “Urinary tract infection, site not specified”, which is infectious, as well as several other non-infectious diseases. We classified each as falling into one of the following groups: "No known infectious agent" (NKIA), where the diagnosis has no infectious component, "Single Known Infectious Agent" (SKIA), which includes codes like J11, where the diagnosis "influenza" is caused by influenza, and finally "Multiple/suspected/non-specific infectious agents" (MSNSIA), for which N39.0 would be an example as numerous infectious agents can cause urinary tract infections, even though N39 at the 3-character level would still be considered NKIA.

Supplemental Dataset 2 | Full pathogen-disease results across both UK Biobank and TriNetX.

This document contains the results from our International Classification of Diseases 10^th^ revision (ICD10) analysis with association test results for all 8,616 pathogen-disease pair tests.

Supplemental Dataset 3 | Full pathogen-Phecode results across both UKB and TNX.

This document contains the results from the Phecode analysis, where Phecodes replaced International Classification of Diseases 10^th^ revision (ICD10) codes as the endpoint but otherwise is identical to our ICD10-based analysis, with association test results for all 18,934 pathogen-Phecode pair tests.

Supplemental Dataset 4 | NCBI GEO RNA-seq datasets used for VIRTUS analyses. This document contains information on NCBI GEO data sets used for our VIRTUS analyses. The first sheet, “SLE”, includes information on the six systemic lupus erythematosus (SLE) case/control RNA-seq data sets performed in blood and B cell subsets used for the Epstein-Barr virus (EBV)-SLE VIRTUS analysis. The second sheet, “UC”, shows the same information for the seven ulcerative colitis (UC) case/control RNA-seq data sets examined in the cytomegalovirus (CMV)-UC VIRTUS analysis.

Supplemental Dataset 5 | RELI Results for EBV-SLE and CMV-UC.

This document shows the RELI results for both the set of differentially expressed genes upon either Epstein-Barr virus (EBV) or cytomegalovirus (CMV) infection and those for the random sampling of expressed but unaltered genes upon infection for all datasets tested. The sheet “EBV_SLE” has the results for the EBV-infected gene sets and risk loci for systemic lupus erythematosus (SLE). The sheet “CMV_UC” provides the RELI results for gene sets from CMV-infected cells and GWAS risk loci for ulcerative colitis.

Supplemental Dataset 6 | Top 175 log product frequency ranked results that were manually reviewed to assemble “Tier 2” positive controls.

This document contains the top 175 most co-cited disease-pathogen pairs as determined by a negated form of the log product frequency (LPF), where the closer the LPF value is to zero, the more commonly the disease and pathogen are found to be co-cited rather than cited individually. After manual review, we limited this list to just pairs with a ‘High’ level of evidence, leaving us with 83 “Tier 2” positive controls. The column “Evidence” contains the evidence level classification, “Mechanism” indicates whether there is a known mechanism by which the pathogen is involved with the disease, “Notes” contains notes taken by M.L. while reviewing, and the “Reference” column includes information about which references were used in the filling of the previously mentioned columns.

Supplemental Dataset 7 | Automated PubMed querying and log product frequency calculation results.

This document contains several sheets, including the calculation of the negated form of the log product frequency (LPF) values for all (426 disease x 20 pathogens) 8,520 disease-pathogen pairs (LPF_data sheet). Because the LPF formula (**Formula 2)** contains a log_10_ transform, the LPF value equals positive infinity for any disease-pathogen pairs with zero co-citations. All three sheets contain the PubMed queries run (“pair_query” on “LPF_data” sheet, “dis_query” on “disease_info” sheet, and “path_query” on “path_info” sheet) to get the PubMed IDs (PMIDs) containing either the co-citations or individual citations (“pair_PMIDs” column for co-citations and “dis_PMIDs” or “path_PMIDs” columns for citations for each disease or pathogen by itself), and the total count of PMIDs found for a co-citation or individual citations (“pair_count” column for number of co-citations, “dis_count” and “path_count” for number of citations for each disease or pathogen individually).

Supplemental Dataset 8 | GWAS risk loci and RNA-seq data sets used for RELI analyses.

This document contains both the RNA-seq data sets (“Gene Sets” sheet) and the GWAS risk loci downloaded from The GWAS Catalog (“GWAS loci” sheet) that were used as inputs for our RELI analyses. Note that GSE99454, a CMV-infection experiment, used two different cell types (lung fibroblast and retinal pigment epithelium), which were analyzed separately. Also, GWAS loci with "-null" in the name indicate the disease-matched (number of SNPs) list of random independent SNPs sampled from snp151Common.

